# Automated Seizure Classification Using Multimodal Large Language Models

**DOI:** 10.1101/2025.10.07.25337538

**Authors:** Lina Zhang, Richard Jiang, Tonmoy Monsoor, Jessica N. Pasqua, Colin M. McCrimmon, Prateik Sinha, Kartik Sharma, Muayad Alzuabi, Victor Morales, Hailey M. Miranda, Chaya Manjeshwar, Vwani Roychowdhury, Rajarshi Mazumder

## Abstract

**Objective:** Accurately distinguishing between epileptic seizures (ES) and nonepileptic seizures (NES) is a significant clinical challenge that typically requires resource-intensive inpatient video-EEG monitoring. Here, we developed a novel Multimodal Large Language Models (MLLMs)-based method for automated extraction of semiological features from videos of seizure events, and subsequently, classified the events as ES or NES.

**Methods:** 90 videos of ES and NES events from 29 patients were obtained from an epilepsy monitoring unit at a large academic hospital. Events were labeled as ES or NES based on expert evaluation of video-EEG recordings and simultaneously annotated with 24 clinically relevant semiological features. We implemented a MLLMs framework that integrates open-source vision-language models (VLMs) and audio-language models (ALMs) to analyze the videos and associated audio tracks and automatically extract these 24 features. The performance of the MLLMs-based feature extraction was evaluated against expert annotations. These features were subsequently used to train several classifiers including K-Nearest Neighbors (KNN), XGBoost, and Deep Factorization Machine, to differentiate ES from NES. Model performance was evaluated using leave-one-patient-out (LOPO) cross-validation.

**Results:** Using KNN, expert-annotated semiological features achieved precision 0.97, recall 0.97, F1-score 0.97, and AUC 0.99, establishing an upper bound on ES/NES classification performance. The MLLMs pipeline achieved an overall mean recall of 0.71, mean accuracy of 0.58, and a mean F1-score of 0.51 for semiological feature extraction compared to expert annotations. The best performing KNN model (k=7) using MLLMs-extracted features achieved a precision of 0.77, recall of 0.76, F1-score of 0.76, and AUC of 0.76 in classifying ES versus NES; correctly identifying 68 out of 90 events.

**Conclusion:** We demonstrate the feasibility of using MLLMs to automatically extract clinically relevant semiological features from seizure videos and classify ES versus NES. MLLMs-based feature extraction and classification offer a promising clinically interpretable approach to aid diagnosis of epilepsy using videos.

## 1. Introduction

Video recordings of seizure episodes provide a rich source of semiological information, particularly by capturing objective clinical manifestations of behavioral and motor changes ^1^. Accurate identification of these characteristic semiological features present during an ictal event is not only critical for establishing an epilepsy diagnosis and localizing the seizure onset zone but is also essential for differentiating epileptic seizures from non-epileptic paroxysmal events or movement disorders that may mimic epilepsy ^2^. The objective semiological features observed during a seizure form the basic framework for correctly distinguishing epileptic seizures from other non-epileptic paroxysmal events ^3^. Thus, semiology plays a fundamental role in defining and classifying seizure and epilepsy syndromes. Recognizing the importance of semiological descriptors, *the ILAE Commission on Diagnostic Methods involving the EEG Task Force* has developed a standardized glossary of descriptors for commonly occurring semiological motifs or clinical features to provide a unified language to describe subjective experiences, behavioral manifestations and motor phenomena occurring during a seizure ^4^.

In this paper, we take advantage of developments in the field of large language models (LLMs) and their multimodal extensions in vision-language models (VLMs) such as GPT-4 Vision ^5^, InternVL ^6^ and audio-language models (ALMs) to identify semiological features from videos of seizure events. Given semiological features of a seizure could be expressed in descriptive natural language; we explore the use of vision-language model to identify clinically relevant features and automate the diagnosis of seizure events. Figure 1 outlines the architecture of a vision language model and its use in identifying seizures from videos. These multimodal approaches incorporate and combine visual and audio perception with natural language understanding to provide higher level reasoning and make inferences about the events occurring in the videos ^7^. VLMs enable natural-language interpretation of movements observed in video frames, similar to the process used by clinicians.

**Figure 1:**
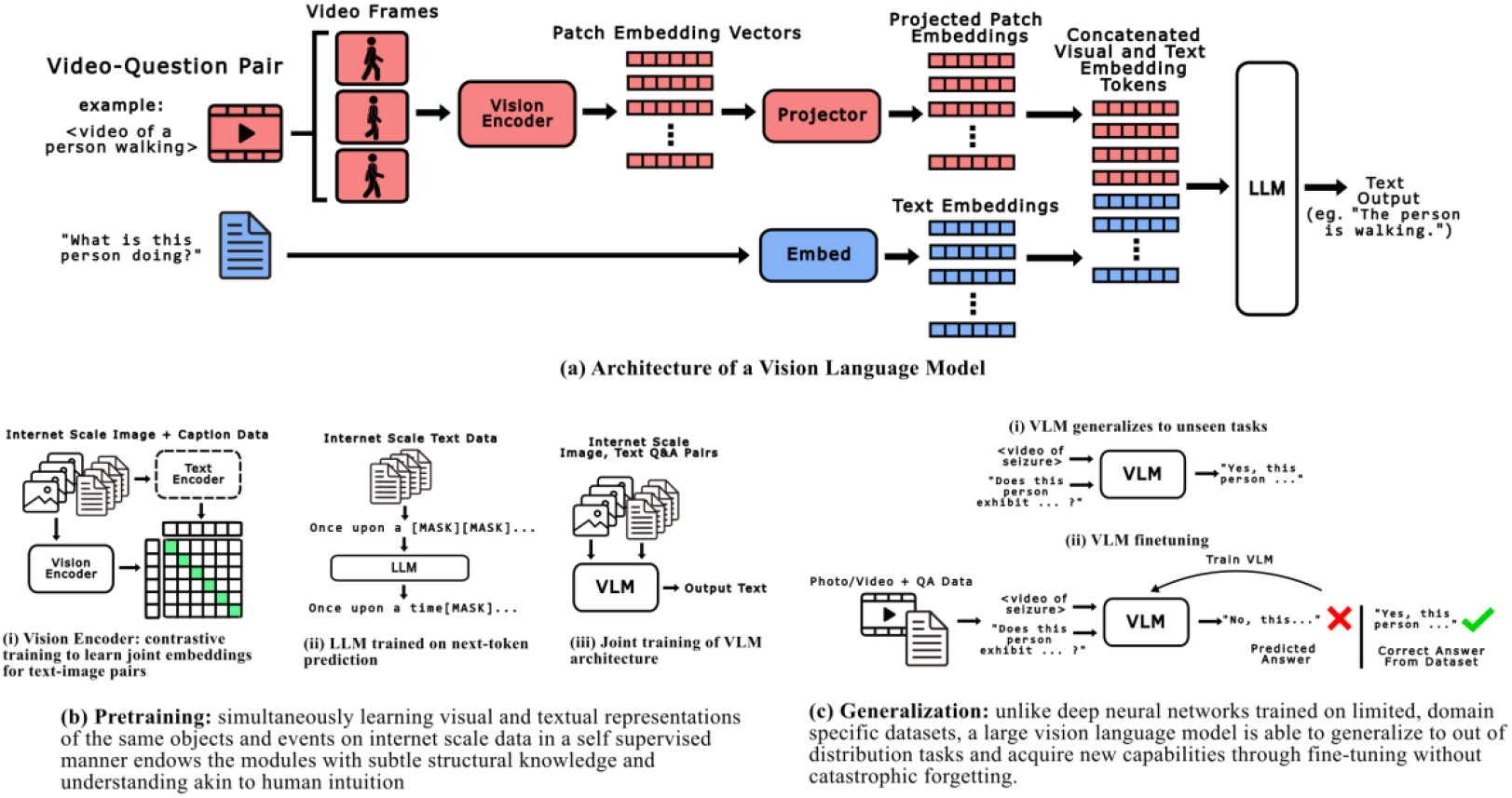
Schematic diagram outlines the architecture of a Vision Language Model and its use in identifying seizures.

Previous approaches to automated video-based seizure analysis often employed a pipeline in which object detection CNNs localized patients’ body parts, pose estimation extracted key points or skeletons, and motion representations such as optical flow captured dynamics ^8^; these features were then fed into deep models (e.g., CNN-LSTM, transformers) to classify events as epileptic seizures (ES), non-epileptic events (NES), or specific seizure types ^9^. Several studies have developed vision-based methods to automate and quantify semiology using videos by performing two primary tasks: a) seizure onset detection and b) seizure classification ^10^. While automated seizure classifications achieved moderate to high accuracy, identifying the semiological features from seizure videos or the sequences of semiological motifs occurring during seizures remains a challenge. Furthermore, the current vision-based seizure classification methods lack clinical interpretability. How and what semiological features are being recognized by the models remain opaque; thus, limiting their clinical utility to further meaningfully characterize seizure events in understandable human language within the framework developed by the ILAE.

We hypothesize that Multimodal Large Language Models (MLLMs) can serve as “virtual neurologists,” observing a seizure through video and audio inputs and identifying key clinical features in a manner analogous to expert human assessment. In this study, we propose the following aims: 1) develop a novel MLLMs-based framework to automatically extract clinically established semiological features from seizure videos; and 2) classify videos as ES vs NES based on the MLLMs-extracted clinical semiology.

## 2. Methods

### 2.1 Dataset and data acquisition

90 video-recorded seizure events from 29 consecutive patients aged >18 years, who underwent video-EEG recording in the epilepsy monitoring unit at the UCLA Medical Center between 2019 and 2023 were included in the study. Videos were selected if they met the following inclusion criteria: 1) epileptic seizures with motor seizure semiology, including a) focal seizure to bilateral tonic-clinic seizures; b) generalized tonic-clonic, c) tonic, d) clonic, and/or e) myoclonic; 2) non-epileptic events with motor phenomena; 3) the whole body of the patient was seen in the video without obstruction. Videos of patients where movement of the body was obscured by blankets were excluded. Definitive diagnosis for epileptic seizures (ES) and non-epileptic seizures (NES) was made using video-EEG recording (conventional 10-20 montage recorded on a Nihon Koden system), which were confirmed by review of medical records and review of the video-EEG reports. A total of 45 ES videos and 45 NES videos were included for this study. All videos were recorded using fixed overhead SONY EP 580 camera at a typical resolution of 1080 X1920 pixels at 30 fps and audio microphones in the epilepsy monitoring unit. Audio was captured at 44.1 kHz mono. Each recording included pre-ictal, ictal, and post-ictal phases with varying durations. Information from concurrent EEG recordings for the videos were used to assign a definitive diagnosis of epileptic seizure (ES) or nonepileptic seizure (NES) and not used in training or testing of the models in this study.

### 2.2 Automated multimodal large language models (MLLMs) based feature extraction and seizure classification

Our workflow comprises three sequential stages, shown in the three rows of **Fig. 2**.

**Figure 2:**
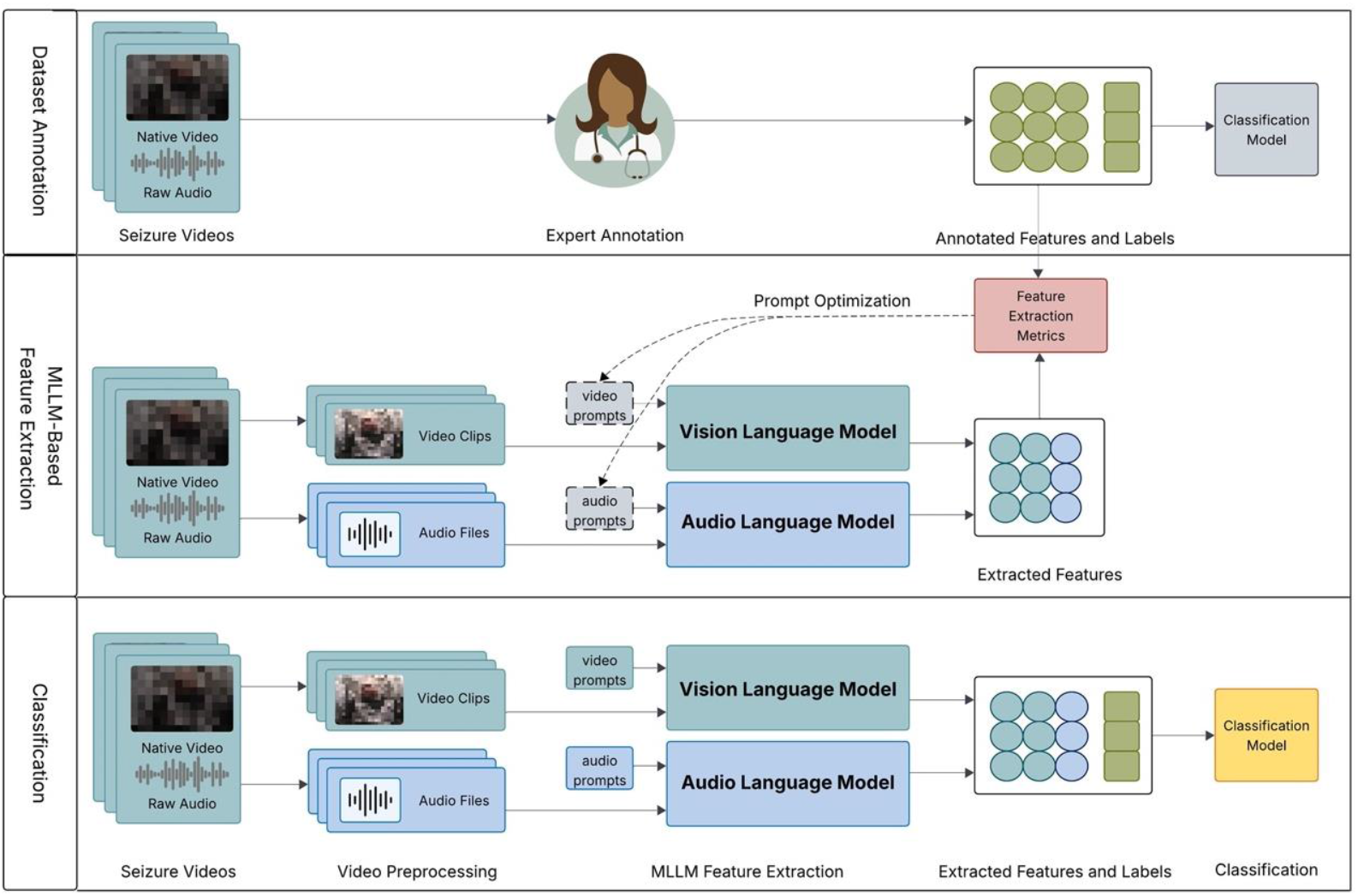
Overall pipeline of seizure classification using Multimodal Large Language Models (MLLMs)

#### A. Human expert dataset annotation (Fig. 2, top row)

Video clips of seizure events (both ES/NES) were reviewed by epileptologists and annotated for the presence or absence of predefined semiological features.

#### B. Automated MLLMs-based feature extraction (Fig. 2, middle row)

Each video clip was temporally segmented and, each segment, accompanied by a feature-focused video prompt, was inputted to an open-source Vision-Language Model (InternVL 3-78B ^6^). Synchronized audio track, guided by matched audio prompts, was processed by an open-source Audio-Language Model (Qwen-2 Audio ^11^) to yield a feature vector. Both the video and audio prompts were iteratively optimized. The two modality-specific vectors were subsequently linked to comprehensive multimodal representations which were compared with expert annotations for feature extraction performance metrics.

#### C. Classification model training and performance measures (Fig. 2 bottom row)

The extracted feature vector along with expert annotated ES/NES labels were used as the training and testing dataset for classification models. The model performances were evaluated using Leave-one-patient-out (LOPO) cross validation.

### 2.3 Human expert dataset annotation and classification

Video clips of seizure events (both ES/NES) were reviewed by two epileptologists (RM, JP) and annotated for the presence or absence of predefined sets of semiological features (Supplement Table1). Concurrent EEG recordings of these seizure episodes were also used to confirm a diagnosis of epileptic seizure (ES) or nonepileptic seizure (NES). The pre-defined semiological features (dichotomized as presence or absence in the video clips) were chosen based on commonly observed signs in epileptic motor seizures and nonepileptic seizures, drawing from the prior literature and clinical experience of the epileptologists.

The expert annotations of semiological features served two purposes: (1) as “ground-truth” labels to evaluate the feature-extraction ability of our MLLMs, and (2) to estimate the maximum achievable classification performance, assuming feature extraction by epileptologists as the “best-case” scenario. The semiological features reflect known factors that differentiates between ES and NES. We constructed a 24-dimensional semiological feature vector where the experts annotated a presence or an absence of the features for each video. The feature vectors along with the video labels were then used to train and test a classification model to obtain a hypothetical upper bound on the ES vs NES classification performance.

### 2.4 Automated MLLMs-based feature extraction

#### 2.4.1 Video preprocessing

Original videos were converted from a M2T to MP4 format. Given current VLMs maximum input token limitation and their markedly better understanding capabilities over short video segments ^12, 7^, we segmented videos into 30-second clips. Each segment was then processed by sampling 64 frames, a common VLMs input capacity, ensuring a frame rate of at least 2 FPS (64 frames/30s). This frame rate was aimed to capture essential motion dynamics of seizure semiologies, as rates below 2 FPS can lead to critical visual information loss. Finally, a 5-second overlap between consecutive segments was introduced to maintain temporal continuity and prevent diagnostically relevant features from being fragmented across segment boundaries, thereby preserving their integrity for feature extraction.

#### 2.4.2 Extract visual features using vision-language model (VLM)

Semiological feature extraction was performed with three state-of-the-art, open-source vision-language models (VLMs): InternVL 2.5-78B ^13^, InternVL 3-78B ^6^, and Qwen2.5-VL-32B ^14^. All model inference was conducted locally on a workstation equipped with four NVIDIA RTX 6000 GPUs. We used a direct question-answering (QA) approach on 30-second video segments. For each segment, the VLMs were input with a focused, binary (yes/no) prompt targeting a single clinical semiological feature. For instance, to detect versive head movement, the prompt asked: “Does the patient forcibly rotate their head to one side during the event?” The models were instructed to return a structured JSON object in the format {“answer”: “yes|no”, “reason”: “justification”}. The answer field represents the feature extraction result, and the reason field provides a textual justification that can be reviewed to verify whether the VLM correctly interpreted the prompt and to identify the visual evidence it used to produce the answer. Segment-level outputs were then consolidated into comprehensive video-level features according to predefined rules: (i) sex was determined by majority vote across segments; (ii) occur during sleep was taken from the first segment containing ictal activity; and (iii) all other semiological features were considered present if detected in at least one segment (“any-yes” rule). As depicted in the middle row of Figure 2, VLM extracted features were compared with expert annotations. We inspected error cases, analyzed the VLM generated reasons, and iteratively optimized prompts. Concise, unambiguous prompts proved most effective, and the final versions are listed in Supplementary Material Table 1.

**Table 1.**
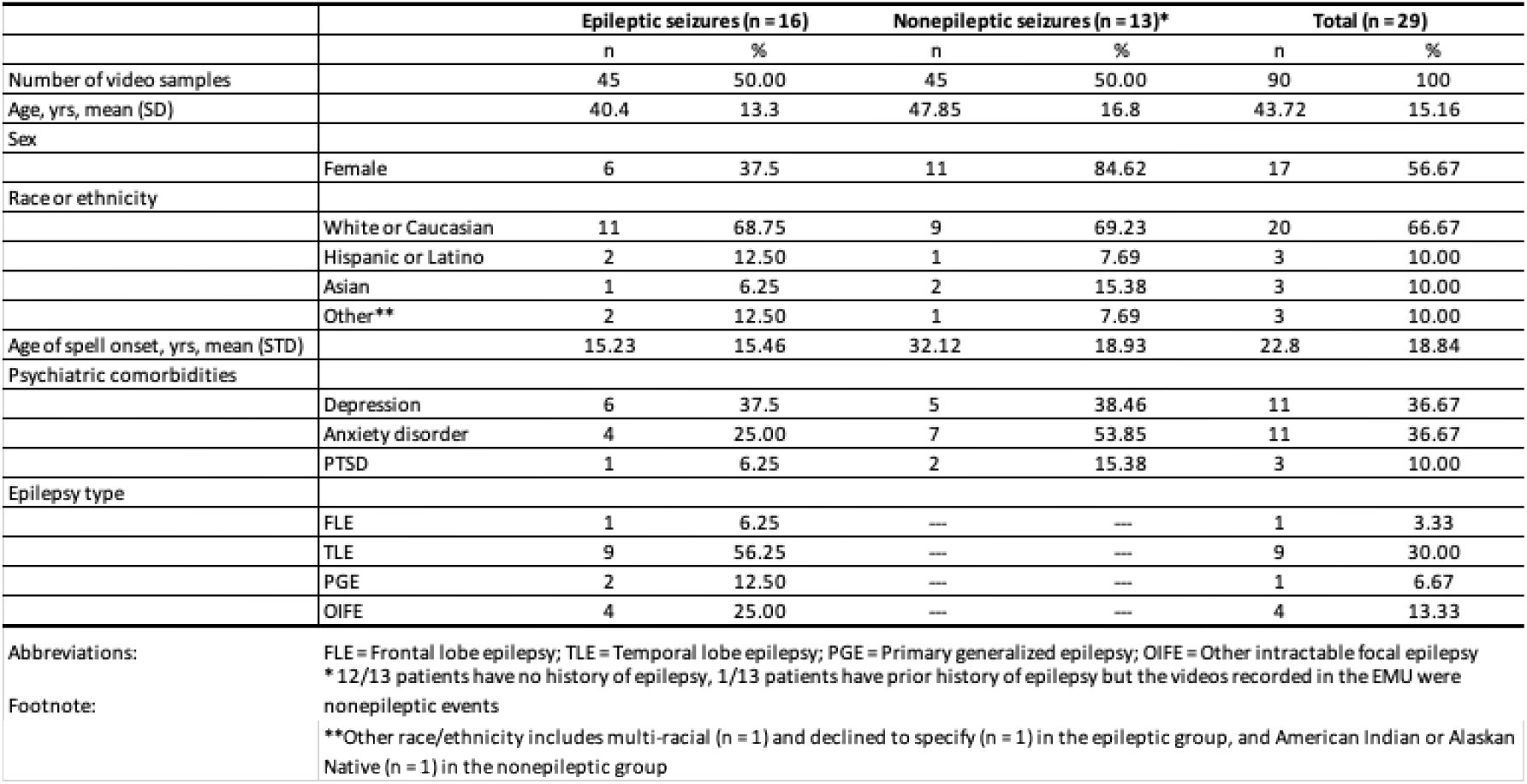
Demographic characteristics of participants with epileptic and nonepileptic seizures.

#### 2.4.3 Extract auditory features using audio-language model (ALM)

To complement the visual pipeline, we analyzed the entire audio track of each seizure episode with the open source ALM Qwen‐2 Audio ^11^. Unlike the video stream, the audio signal was not segmented: the full waveform corresponding to the recorded event was fed directly into the model, ensuring that all vocal phenomena were captured in one pass. Two auditory features were queried via separate prompts (see Supplementary Material Table 1 for prompts):

- **Ictal vocalization** – presence of any cry, groan, moan, or scream during the seizure.
- **Verbal responsiveness** – intelligible speech produced by the patient in response to caregiver’s or provider’s questioning at bedside. Here we recognize **three outcomes**: “yes” (patient responds), “no” (question asked but no response), or **“NA”** (no question asked, therefore responsiveness cannot be assessed).

Because each audio file was analyzed as a single unit, the ALM output directly yielded the video‐level results for ictal vocalization and verbal responsiveness, therefore no post‐processing aggregation was required.

### 2.5 Classification model training and performance measures

#### 2.5.1 Feature vector and label construction

For each seizure video, we constructed a 24-dimensional feature vector where each entry of the vector corresponds to a semiological feature. The 24-dimensional feature vector were populated using the textual values (‘yes’,’no’,’N/A’) of the semiological features automatically extracted by the MLLMs-based framework. Finally, each ES video was assigned a class label of 1 and each NES video was assigned a class label of 0.

#### 2.5.2 Classification model

We explored several classifiers, from simple to more complex, to comprehensively evaluate classification metrics: (i) K-Nearest Neighbors (KNN) classifies an event by a majority vote of the k most similar past events in the training set. This method is simple and can perform well if the features cluster distinctly by class. (ii) XGBoost (XGB) ^15^ is well-suited for small-to-medium tabular datasets and can capture nonlinear feature interactions. Importantly, it provides feature importance scores and tree-based explanations. (iii) Deep Factorization Machine (DeepFM) ^16^ is a neural network that combines the strengths of factorization machines for modeling low-order feature interactions and deep neural networks for capturing high-order feature interactions. Considering our dataset size of 90 samples, deep learning models like DeepFM are prone to overfitting. Furthermore, deep learning networks often act as “black boxes,” making it challenging to derive feature importance. Thus, we included DeepFM primarily for completeness in our comparison and did not focus on extensive optimization for this model.

#### 2.5.3 Classification model training and testing

All classification models were trained and evaluated using a Leave-One-Patient-Out (LOPO) cross-validation scheme. LOPO is a stringent evaluation where we hold out all events from that patient as the test set, training the model on events from all other patients in each iteration. This evaluates how the model generalizes to a completely new patient, since we ultimately want to classify events from patients the model hasn’t seen before. We report the performance across all LOPO folds (i.e., patients) as the result.

#### 2.5.4 Classification model performance metrics

To calculate performance metrics, predictions from all LOPO folds were first aggregated. The overall **precision, recall (sensitivity), F1-score, and ROC AUC** were then computed from these aggregated results. Precision indicates how many events the model predicted as “epileptic” were truly epileptic. Recall (sensitivity) indicates how many true epileptic seizures were detected. The F1-score is the harmonic mean of precision and recall, summarizing overall classification effectiveness. The AUC measures the model’s ability to discriminate classes across all thresholds, providing a threshold-independent metric.

#### 2.5.5 Semiological feature importance analysis

We examined semiological feature importance **for KNN** using permutation importance measure, which measures the decrease in model performance when the values of a feature are randomly shuffled and indicates how much each feature contributes to reducing classification error. We averaged these importance scores across the LOPO folds to see which clinical features were most useful to the model and ranked them based on the average importance scores. We report the top 10 MLLMs extracted semiological features.

## 3. Results

### 3.1 Demographic characteristics

29 participants (16 ES and 13 NES) were included in the study. 45 videos from each group of ES and NES were included. The mean cohort age was 43.7 years (SD = 15.2), with a predominance of females (56.7%). The mean age of spell onset was 22.8 years (SD = 18.8), which was lower in ES (15.2 ± 15.5 years) as compared with NES (32.1 ± 18.9 years). Psychiatric comorbidities were common in both ES and NES groups, most frequently depression and anxiety (each 36.7%), followed by PTSD (10%). Among those with ES, the most frequent epilepsy type was temporal lobe epilepsy (30%), followed by other intractable focal epilepsies (13%), primary generalized epilepsy (7%), and frontal lobe epilepsy (3%).

### 3.2 ES vs NES classification based on semiological features annotated by experts

Using 24 clinical features identified and annotated by experts, the performance of the classification model to distinguish between ES and NES was as following: F1 Score (0.95), Accuracy (0.88), Precision (0.96), Recall (0.95) [Fig. 3C]. Furthermore, the Area Under the ROC Curve (AUC) was 0.99 [Fig. 3D] The 3D PCA representation of the 24 clinical semiological features [Fig. 3A] demonstrates well-defined clustering, with a clear separation between ES and NES. Collectively, these results demonstrate that the KNN model based on expert-defined features, is highly effective at differentiating between ES and NES.

**Figure 3:**
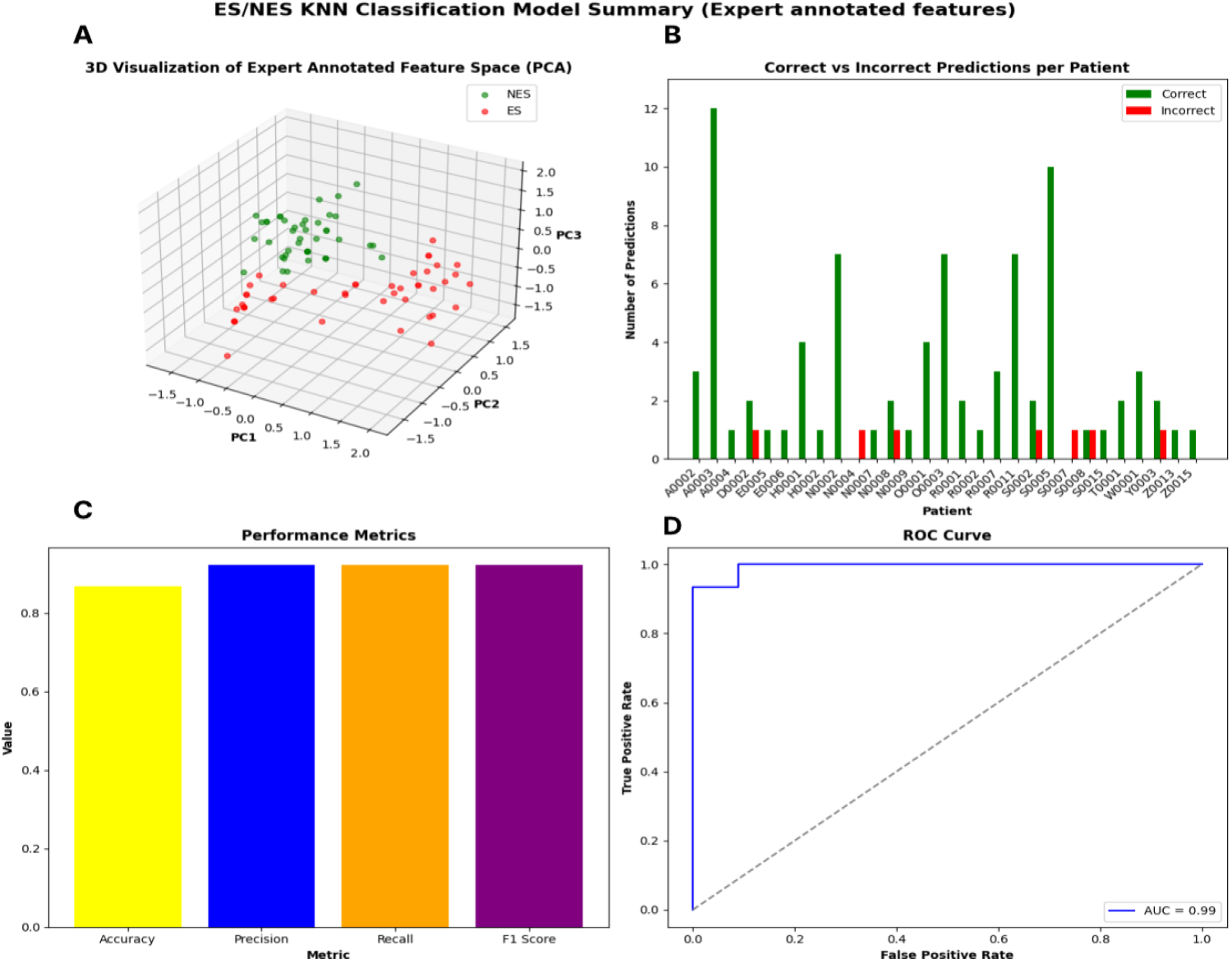
Performance evaluation of the ES/NES KNN classifier based on semiological features annotated by experts using LOPO CV: The 24 clinical semiological features as identified by epileptologists distinguishes between Epileptic Seizures (ES) and Non-Epileptic Seizures (NES) **(A)** 3D Principal Component Analysis (PCA) visualization, demonstrating clear separation in the semiological feature between ES (red) and NES (green) events, suggesting the semiological features chosen by experts are highly discriminative. **(B)** The model’s predictive accuracy on a per-patient basis, showing high number of correct classifications (green bars) across most individuals with few incorrect predictions (red bars). **(C)** Overall performance metrics, with the model achieving Accuracy of 0.88 (yellow), Precision of 0.96 (blue), Recall of 0.95 (orange), and F1 Score of 0.95 (purple). **(D)** Receiver Operating Characteristic (ROC) curve yielding an Area Under the Curve (AUC) of 0.99.

### 3.3 MLLMs-based automated semiological feature extraction from videos

Next, we evaluated the performance of Multimodal Large Language Model’s (MLLMs; VLM+ALM) semiological feature extraction against expert annotations. Table 2 shows the performance achieved for each of the 23 semiological features (duration is not included). The MLLMs model overall demonstrated notable efficacy, achieving a mean recall of 0.71, mean accuracy of 0.58, mean F1 of 0.51 and mean precision of 0.43.

**Table 2.** MLLMs Feature Extraction Metrics: Optimal accuracy and recall per feature from tested MLLMs. “Close eyes” and “face pulling” were best extracted by InternVL 2.5 (78B); all other features by InternVL 3 (78B).

|  | gender | occur_during_sleep | head_turning | blank_stare | close_eyes | eye_blinking |
| --- | --- | --- | --- | --- | --- | --- |
| Accuracy | 0.94 | 0.70 | 0.33 | 0.62 | 0.48 | 0.67 |
| Precision | 0.95 | 0.55 | 0.22 | 0.57 | 0.31 | 0.22 |
| Recall | 0.97 | 0.94 | 0.94 | 0.54 | 0.78 | 0.43 |
| F1 Score | 0.96 | 0.70 | 0.36 | 0.55 | 0.44 | 0.29 |
|  | facepulling | face_twitching | tonic | clonic | arm_flexion | arm_straightening |
| Accuracy | 0.41 | 0.38 | 0.66 | 0.72 | 0.68 | 0.54 |
| Precision | 0.32 | 0.38 | 0.46 | 0.40 | 0.74 | 0.38 |
| Recall | 0.76 | 1.00 | 0.41 | 1.00 | 0.67 | 0.78 |
| F1 Score | 0.45 | 0.55 | 0.44 | 0.58 | 0.70 | 0.51 |
|  | figure4 | oral_automatizms | limb_automatizms | pelvic_thrusting | full_body_shaking | asynchronous_movement |
| Accuracy | 0.60 | 0.42 | 0.41 | 0.50 | 0.66 | 0.60 |
| Precision | 0.13 | 0.33 | 0.19 | 0.22 | 0.47 | 0.54 |
| Recall | 0.62 | 0.65 | 0.48 | 0.80 | 0.89 | 0.85 |
| F1 Score | 0.22 | 0.43 | 0.27 | 0.35 | 0.62 | 0.66 |
|  | motor_pattern | limb_movement_pattern | hands_move_simultaneously | verbal_responsiveness | ictal_vocalization | mean |
| Accuracy | 0.69 | 0.60 | 0.64 | 0.51 | 0.58 | 0.58 |
| Precision | 0.58 | 0.54 | 0.32 | 0.58 | 0.59 | 0.43 |
| Recall | 0.55 | 0.60 | 0.29 | 0.51 | 0.94 | 0.71 |
| F1 Score | 0.56 | 0.55 | 0.30 | 0.51 | 0.72 | 0.51 |

### 3.4 ES vs NES classification using MLLMs extracted features

#### 3.4.1. Comparison of performance of different classification models

Table 3 compares the performance of various classification models to distinguish between ES and NES based on semiological features extracted by MLLMs. The KNN model, when utilizing the full set of MLLMs-extracted features including duration of the seizure events, provides an accuracy of 0.743, precision of 0.764, recall of 0.756, a F1 score of 0.754, and an AUC of 0.761. The XGBoost model also achieves robust results with an accuracy, precision, recall, and F1 score of 0.700, and an AUC of 0.757. In contrast, the DeepFM model exhibits lower performance (Accuracy of 0.549, AUC of 0.662).

**Table 3.** ES/NES Classification Model Performance Comparison using MLLMs extracted features.

| Model | Accuracy | Precision | Recall | F1 score | AUC |
| --- | --- | --- | --- | --- | --- |
| KNN | 0.743 | 0.764 | 0.756 | 0.754 | 0.761 |
| XGBoost | 0.700 | 0.701 | 0.700 | 0.700 | 0.757 |
| DeepFM | 0.549 | 0.656 | 0.644 | 0.638 | 0.662 |

#### 3.4.2 Performance evaluation of the ES/NES KNN classifier

The optimal performance for the KNN model, identified with k=7 [Fig. 4A], provided an Area Under the ROC Curve (AUC) of 0.76 [Fig. 4D]. The confusion matrix [Fig. 4C] based on the KNN classifier using the optimal k=7, shows that the MLLMs, correctly identifies a substantial majority of ES cases, specifically 38 out of 45. This translates to a sensitivity of approximately 84.4% for ES. Simultaneously, the model correctly classified 30 out of 45 NES cases (representing a specificity of 66.7%).

**Figure 4:**
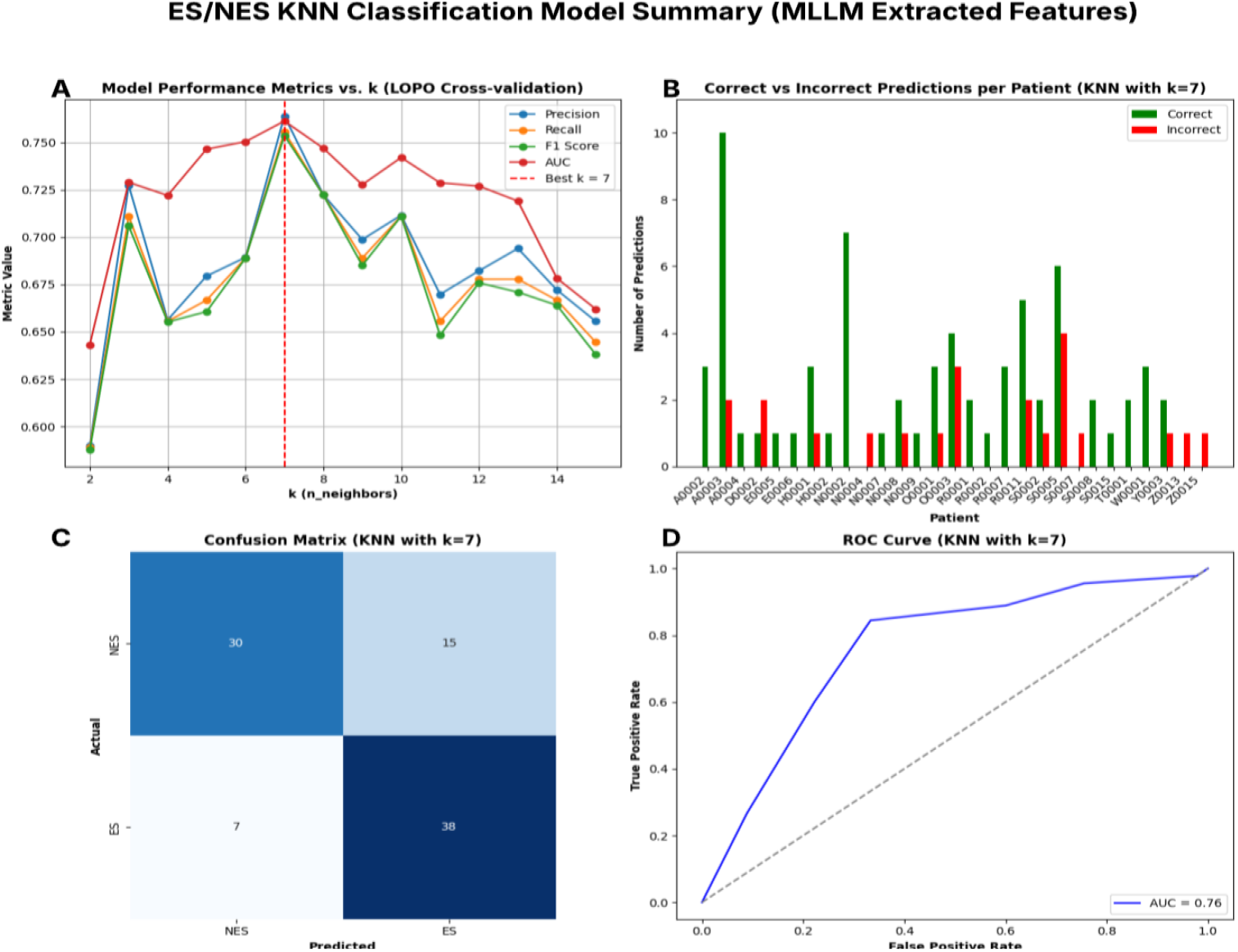
Performance Evaluation of the ES/NES KNN classifier using MLLMs extracted features and LOPO CV: (A) The performance of the KNN model evaluated using LOPO cross-validation. The x-axis represents the value of ‘k’ (from 2 to 15), and the y-axis represents the metric score. Four key performance metrics are plotted: Precision (blue), Recall (orange), F1 Score (green), and Area Under the Curve (AUC, red). k=7 as the optimal value was chosen based on maximizing the AUC, indicating the best overall discriminative ability found during cross-validation. (B) Prediction performance of the optimized KNN model (with k=7) on an individual patient basis. (C) Confusion matrix for the KNN classifier using the optimal k=7. (D) Receiver Operating Characteristic (ROC) curve for the KNN model with k=7, the calculated Area Under the Curve (AUC) is approximately 0.76.

### 3.5 Top 10 MLLMs extracted semiological features retain sufficient discriminative power despite feature reduction

Using permutation feature importance, we identified the top 10 MLLMs annotated features which distinguished between ES and NES [Fig. 5]. The K-Nearest Neighbors (KNN) model, when utilizing only these top 10 features (including prominent ones like “arm flexion”, “limb movements pattern”,” motor pattern”, etc.), achieved an Area Under the ROC Curve (AUC) of and an F1 Score of 0.70. While this represents a slight decrease from the 0.76 AUC and F1 Score obtained from using all 24 MLLMs annotated features, the performance with the reduced feature set maintained sufficient discriminative power. “Duration” is identified as the most impactful feature, followed by “arm flexion,” “limb movements pattern,” and “motor pattern”. Other top features that reliably distinguished between ES and NES include “arm straightening,” “full body shaking,” “verbal responsiveness”, “close eyes”,”occur during sleep”, and “pelvic thrusting”. Identification of key features for the classification task enables an interpretable and clinically relevant diagnosis.

**Figure 5:**
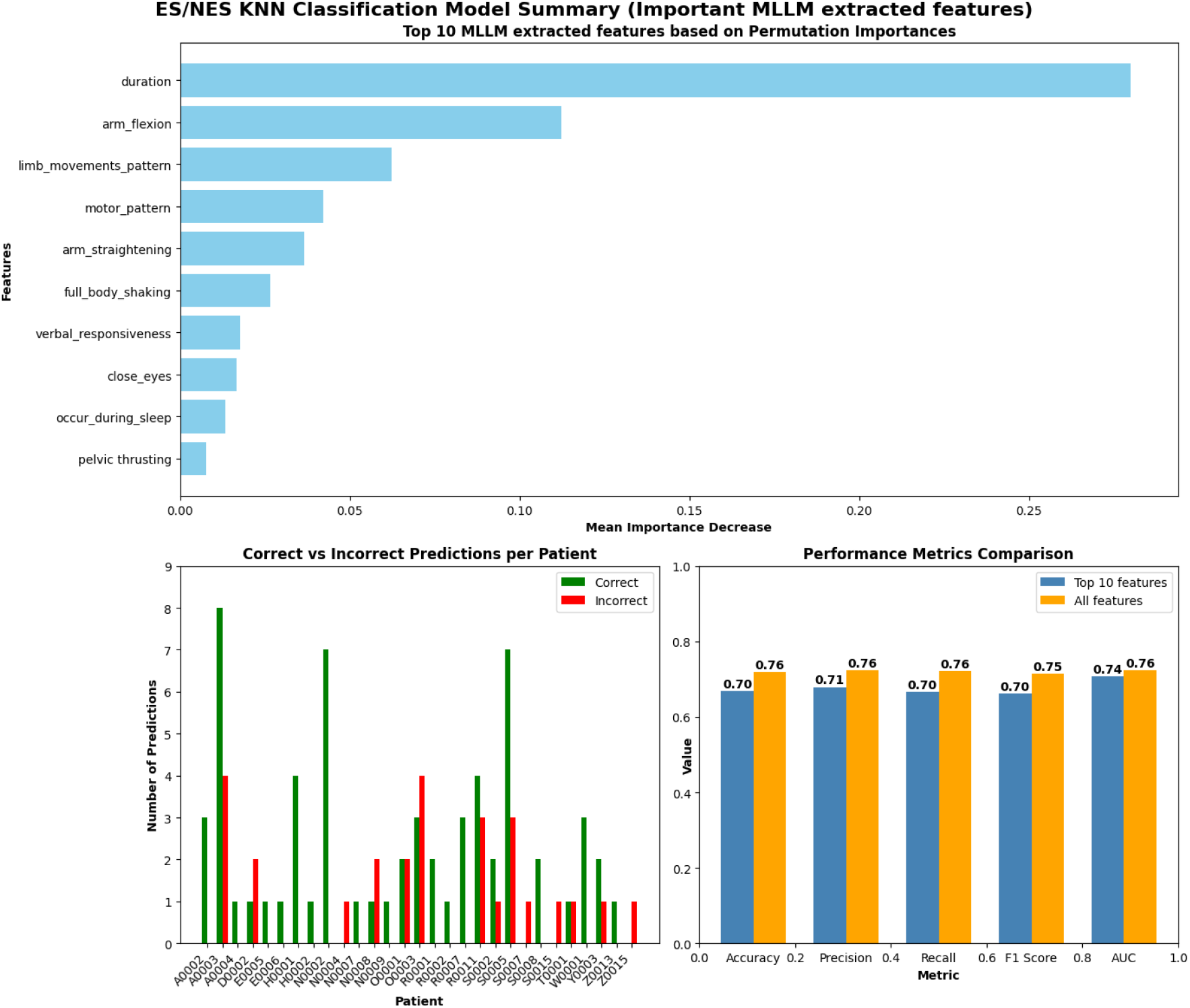
K-Nearest Neighbors (KNN) Classification Model Performance using Important Multimodal Large Language Model (MLLMs) Extracted Features: **(A)** The ten most significant features extracted by the MLLMs, as determined by their mean importance decrease in the KNN model’s predictive performance when their values are permuted. The length of each bar corresponds to the calculated importance, indicating the extent to which the model’s performance degrades when that feature’s information is randomized. **(B)** Patient-specific prediction outcomes of the KNN model when trained using the top 10 MLLMs-extracted features identified in Panel A. Green bars represent correct classifications, while red bars denote incorrect classifications. **(C)** Comparison of key performance metrics—Accuracy, Precision, Recall, F1 Score, and Area Under the ROC Curve (AUC)—for the KNN model with top 10 most important MLLMs-extracted features (blue bars) and a model using all MLLMs-extracted features (orange bars).

### 3.6 Explainability of the multimodal large language model (MLLMs) in distinguishing between epileptic seizures (ES) and non-epileptic seizures (NES)

Table 4 showcases the explainability of the MLLMs in distinguishing between Epileptic Seizures (ES) and Non-Epileptic Seizures (NES). Instead of acting as a black box, the MLLMs identifies and lists specific semiological features extracted from patient videos that allows the classification of ES and NES. As an example, for patient N0002 with ES, the MLLMs identified features such as “arm flexion,” “oral automatisms”, “ictal vocalization”, “face twitching”, “occur during sleep”, and “close eyes”. Subsequently, for patient O0003 with NES, the MLLMs extracted features like “pelvic thrusting”, “eye blinking”, and “full body shaking”.By detailing these specific features, the MLLMs provides transparency for the characteristics it deems significant for differentiating between ES and NES. This provides a method for clinical interpretation and improves explainability.

**Table 4.** Patient specific feature extractions by MLLMs.

| Video Name | Features extracted by MLLMs |
| --- | --- |
| N0002 Episode 1 (ES) | arm flexion,oral automatisms,ictal vocalization,face twitching,occur during sleep,close eyes |
| N0002 Episode 2 (ES) | Arm straightening,head turning,ictal vocalization,face twitching |
| O0003 Episode 1 (NES) | pelvic thrusting,eye blinking, full body shaking |

### 3.7. Multimodal feature integration

We incorporated both video and audio information in our pipeline, hypothesizing that this combination would yield a more complete and robust depiction of each seizure event than either modality alone. To test this assumption, we performed an ablation study that removed auditory features; the F1 score subsequently fell from 0.75 to 0.71 and the AUC dropped from 0.76 to 0.74, as detailed in Supplementary Material Table 2. The performance loss confirms that a vision only approach misses critical diagnostic information available in the acoustic domain.

## 4. Discussion

In this study, we developed an automated multimodal AI pipeline that extracts clinically meaningful visual and auditory semiological features from patient videos and classifies each event as an ES or a NES. We found that features extracted using an MLLMs reliably distinguished between ES and NES through automated video-audio analysis. While the performance metrics achieved with MLLMs-extracted features could not fully match those obtained by experts, the results show that features derived from an MLLMs, even with minimal human intervention via prompting, could reliably extract the clinically meaningful characteristics necessary for a complex seizure classification task. Our current classifier achieved an F1 score of 0.75 and an AUC of 0.76.

To our knowledge, this is among the first studies to apply VLM and ALM to interpret seizure semiology and, subsequently, classify seizure spells as ES or NES. We show that an off-the-shelf vision-language model, without epilepsy-specific training, can assess a seizure video and recognize descriptors such as “eyes closed”, “clonic” or “ictal vocalization”. This finding represents an important step toward the creation of an AI system that functions as a virtual epileptologist which can interpret video features into human language. Earlier work in seizure-video analysis used skeleton-based and frame-based human action recognition frameworks to successfully distinguish ES and NS but did not attempt to identify commonly recognized clinical and semiological signs^17 18, 19^.

Most prior studies focused separately on seizure semiology detection and classification tasks for distinguishing between ES and NES. A comprehensive review details the description of the different deep learning methods and tasks that these studies accomplished. ^10^ A wide array of deep learning techniques have been employed previously, from convolutional neural networks (CNN) to spatio-temporal CNN combined with features extraction with skeleton-based to frame-based human action recognition modalities and human pose estimation.^20, 21^ Our work adds to the current body of work by combining the task of semiology detection and subsequent seizure classification into a single platform. Multimodal large language model was used to transform visual and auditory cues into explicit semantic descriptions, which then applied to classify seizures based on semiology. This mimics the approach a trained epileptologists uses in evaluating seizures.

A key advantage of our two-step pipeline is the transparency of AI decision making. Unlike a monolithic deep network that might simply output “NES” without explanation, our method accompanies each prediction with a rationale based on clinically described features. For example, when the model classifies an event as NES it provides a list of descriptors such as eyes closed, asynchronous limb movements, and pelvic thrusting, which are signs a neurologist would also recognize as characteristic of NES. Feature-importance analysis of the downstream classifier further highlights which semiology cues drive the decision, adding another layer of interpretability. Rather than producing a black box diagnosis, our MLLMs-based framework provides explicit descriptors that map directly onto clinical reasoning.

Analysis of the MLLMs Feature Extraction Metrics (Table 2) reveals that the multimodal model’s performance varied widely across features. Generally, global features that involves several parts of the body were detected most reliably. The VLM consistently recognized large motor phenomena, such as generalized body jerks or sustained limb postures (e.g., tonic/clonic activity, full body shaking, arm flexion/straightening), and clinical attributes such as the patient’s gender or the context of seizure (e.g., sleeping vs. awake). These features yielded relatively high agreement with expert labels. In contrast, focal features involving smaller body parts such as the face or hands in isolation were often missed. The model frequently failed to detect brief eye blinks, partial eye closure, or fine facial movements. Similarly, distinguishing movement patterns with complex temporal evolution, such as differentiating asynchronous flailing from rhythmic clonic jerking, proved challenging. The overall extraction accuracy for these features remained substantially lower than for larger-scale movements. The ALM component extracted two audio features, “ictal vocalization” and “verbal responsiveness”, with mixed reliability. It detected clear vocal sounds (e.g., cries, moans) robustly, achieving a recall of approximately 0.94 for “ictal vocalization”. However, performance on “verbal responsiveness”, which assessed the patient’s response to a caregiver or a medical provider, was modest (accuracy and recall approximately 0.51), primarily attributed to noise or misidentification when multiple voices overlapped.

Some visual features present considerable difficulties even during expert annotation. Camera angles, lighting conditions, blockades or other obstacles sometimes required experts to repeatedly review video frames to determine the presence of subtle features like “eye blinking” or “limb automatisms”. Experts labeled these symptoms only after a comprehensive review of multiple episodes. Such comprehensive contextual reasoning remains beyond the scope of current VLM abilities.

Even with these limitations, we found that the MLLMs-based automated semiological feature detection and seizure classification has great potential for the future of streamlining video-based epilepsy diagnoses. Automated recognition of semiology opens the door to generate narrative reports of a patient’s seizure episodes. In an epilepsy monitoring unit, the MLLMs-based systems could potentially prescreen hours of video-EEG, flagging seizure-like segments with preliminary ES or NES labels and key semiology; so that clinicians review only relevant video information, greatly reducing time spent meticulously combing through normal activity. Particularly, in resource-limited regions, where epilepsy expertise is scarce, this system could extend neurological support remotely through ubiquitous video capture to provide decision assistance to local providers.

Despite the promise of our current work, there are several limitations to acknowledge. First, the dataset size is small (90 events), which restricts the statistical power of our findings and the generalizability of the trained models. We compared several classification models and found that the DeepFM model exhibits lower performance compared to KNN and XGBoost models. This is likely attributable to the small dataset size of only 90 samples, which can lead to overfitting in more complex models like DeepFM, rather than a deficiency in the MLLMs features themselves. Overall, both KNN and XGBoost demonstrated reliable performance. This strongly suggests that the features extracted by the MLLMs are indeed highly discriminative and effective at the seizure classification task. Additionally, the feature extraction errors significantly impacted classification outcomes. Our MLLMs struggled with some subtle or transient visual features causing misclassifications. To address this, one could fine-tune an MLLMs on a curated dataset of seizure videos in the future. Another limitation was understanding or capturing the temporal evolution of seizures. Our approach parceled videos into segments and queried the VLM on short clips. We did not explicitly model the sequence of motor behavior. Epileptologists often consider the evolution of a spell (start, progression, and end) in making a diagnosis. Capturing these temporal patterns could further enhance differentiation. Future models might prompt the VLM to describe the sequence (“first the patient does X, then Y…”). This was beyond our initial scope but represents fertile ground for future work.

### Ethical Considerations

This study was approved by the UCLA IRB. Our pipeline kept everything local behind a firewall using open-source VLMs/ALMs to avoid transmitting and sharing sensitive patient video data.

## 5. Conclusions

Here we present a multimodal AI approach that integrates vision-language and audio-language models to classify seizures from video recordings. Our pipeline successfully extracted key semiological signs and achieved a moderate classification accuracy in distinguishing epileptic seizures from nonepileptic events. This approach shows the potential of large pre-trained MLLMs to assist in clinical decision making by providing interpretable descriptions of patient behaviors. This work lays the groundwork for future development of automated, explainable seizure diagnostics. Future research will focus on refining feature extraction (including temporal dynamics) and validating with larger patient cohorts.

## Data Availability

All data produced in the present study are available upon reasonable request to the authors

## Acknowledgments

This work was supported by funds from Department of Neurology, David Geffen School of Medicine, UCLA. RM was supported by the Fogarty International Center of the National Institutes of Health under Award Number K01TW012178. The content is solely the responsibility of the authors and does not necessarily represent the official views of the National Institutes of Health.

