## Supplemental Table 1, Supplemental Table 2 for "Automated Seizure Classification Using Multimodal Large Language Models"

### **Supplementary Material**

**Table 1 Semiological Features and Corresponding MLLMs Prompts**

| **Metafeature** | **Feature** | **Description/Definition** | **Prompt** |
|---|---|---|---|
| Patient Characteristics | Sex | Male/female | Please identify the gender of the patient in the video. Please answer with "female" or "male". |
| Context of the seizure | occur_during_sleep[1] | Event of interest occurs while patient is asleep, as visualized in the video recording. | Please determine if this seizure event occurs while the patient is asleep. Answer with 'yes' or 'no'. |
| | duration | The length of time for the seizure event. | |
| Epileptic Semiological features | head_turning[2] | Unnatural forced and sustained versive head turn. | Does the patient forcibly or stiffly rotate their head to one side during the event? Answer with 'yes' or 'no'. |
| | blank_stare[3] | Staring with motor arrest. | Does the patient exhibit a blank stare (vacant or unfocused gaze)? Answer with 'yes' or 'no'. |

| | | | |
|---|---|---|---|
| | close_eyes[4] | Eyes are bilaterally closed | Do the patient's eyes remain consistently closed or mostly closed throughout the seizure? Answer with 'yes' or 'no'. |
| | eye_blinking[5] | Repetitive clonic contraction of the eyelids during a seizure. | Does the patient show repeated or rapid blinking of the eyes during the event? Answer with 'yes' or 'no'. |
| | face_pulling[6] | Tonic contraction of the face. | Does the patient's facial expression indicate grimacing or face-pulling movements? Answer with 'yes' or 'no'. |
| | face_twitching[6] | Repetitive clonic contraction of the face (unilateral or bilateral). | Are there small, involuntary twitches or jerks observed on the patient's face? Answer with 'yes' or 'no'. |
| | tonic[7] | Sudden, sustained body stiffness, typically lasting 5 to 20 seconds. | The tonic phase is characterized by sudden, sustained body stiffness, typically lasting 5 to 20 seconds. This rigidity can be generalized, causing limbs to become fixed in an extended or flexed posture and the trunk and back to straighten or arch, or it can be focal, limited to areas like |

| | | | |
|---|---|---|---|
| | | | the face (grimacing), eyes (deviation), or one side of the body. Does this patient show tonic posture? Give an answer with 'yes' or 'no'. |
| | clonic[8] | Rhythmic, jerking muscle contractions involving the limbs, face, and trunk. | Clonic Phase: Rhythmic, jerking muscle contractions involving the limbs, face, and trunk. During this phase, individuals may bite their tongue or froth at the mouth. The episode typically lasts 30–90 seconds, though it can persist longer. The jerking movements gradually slow before ceasing entirely. Clonic movements present as rhythmic, alternating contraction-relaxation cycles (e.g., limb jerking).Does this patient show clonic movement? Answer with 'yes' or 'no'. |
| | arm_flexion[7] | Sustained arm flexion | Does the patient maintain a sustained flexion of the arms at the elbows during the seizure? Answer with 'yes' or 'no'. |
| | arm_straightening[7] | Sustained arm stiffening in an extended position. | Does the patient straighten or stiffen their arms (extended position)? Answer with 'yes' or 'no'. |

| | | | |
|---|---|---|---|
| | figure4[7] | One upper limb is extended, while the contralateral upper limb is flexed. | Figure 4 refers to a specific posture or movement observed in a patient during a seizure, where one upper limb is extended (typically in a tonic stretch) while the other upper limb is flexed, forming a shape resembling the number "4“. Please check if there is a 'figure 4' posture of the arms at any point. Answer with 'yes' or 'no'. |
| | oral_automatisms[9] | | Does the patient exhibit repetitive mouth or tongue movements such as chewing, lip-smacking, or swallowing? Answer with 'yes' or 'no'. |
| | limb_automatisms[10] | Involuntary, repetitive movements of the mouth or face. | Are there repetitive, purposeless limb movements (e.g., fumbling, picking, patting, cycling) observed? Answer with 'yes' or 'no'. |
| Non-epileptic semiology | pelvic_thrusting[11] | Repetitive forward and backward movements of the pelvis. | Does the patient display any pelvic thrusting movements during the event? Answer with 'yes' or 'no'. |
| Non-epileptic semiology | full_body_shaking[12] | Generalized irregular and asynchronous | Does the patient experience shaking of the entire body— |

| | | | |
|---|---|---|---|
| | | movements involving the entire body. | including arms, legs, torso? Answer with 'yes' or 'no'. |
| Non-epileptic semiology | asynchronous_movement [12] | Different limbs move out of phase with one another. | Do you observe the patient's limbs shake with variable intensity with respect to one another? Answer with 'yes' or 'no'. |
| Temporal sequence of body pose/movements | motor_pattern[13] | | Does the patient show a continuous, intense, non-evolving motor pattern? Answer with 'yes' or 'no'. |
| Temporal sequence of body pose/movements | limb_movements_pattern[14] | | Analyze the patient's upper limb movements in this video. Which of the following best describes the movements?<br>1.Thrashing/Flailing: Significant thrashing and flailing, asynchronous and variable.<br>2.Rhythmic Shaking: Stereotyped, rhythmic clonic shaking, potentially following a tonic phase.<br>3.Neither: The movements do not fit descriptions 1 or 2.<br>Please answer with 1, 2, or 3. Do not include extra text in your output—only the answer. |

| | | | |
|---|---|---|---|
| Temporal sequence of body pose/movements | hands_move_simultaneously[14] | | Do the patient's hands start moving simultaneously? Answer with 'yes' or 'no'. |
| | verbal_responsiveness[15] | | Please determine if the patient is able to respond verbally during the event. Your answer must be one of the following three options. 'yes': The patient gave a verbal response when the doctor spoke to them. 'no': The doctor attempted to communicate, but the patient showed no verbal response. 'NA': No attempt was made by doctors to communicate with the patient. Answer with 'yes', 'no', or 'NA' only. Do not include extra text in your output—only the answer. |
| Audio | ictal_vocalization[16] | | Please check if the patient produces any vocalization (such as groaning, moaning, or screaming) during the event. Answer 'yes' or 'no'. |

**Table 2 Ablation Study - ES/NES KNN Classification Model Performance Comparison without some features:** fill this out

| Model | Accuracy | Precision | Recall | F1 score | AUC |
|---|---|---|---|---|---|
| KNN | 0.743 | 0.764 | 0.756 | 0.754 | 0.761 |
| KNN w/o duration | 0.682 | 0.701 | 0.700 | 0.700 | 0.710 |
| KNN w/o audio | 0.708 | 0.722 | 0.722 | 0.722 | 0.743 |
